# Mapping malaria by sharing spatial information between incidence and prevalence datasets

**DOI:** 10.1101/2020.02.14.20023069

**Authors:** Tim C.D. Lucas, Anita K. Nandi, Elisabeth G. Chestnutt, Katherine A. Twohig, Suzanne H. Keddie, Emma L. Collins, Rosalind E. Howes, Michele Nguyen, Susan F. Rumisha, Andre Python, Rohan Arambepola, Amelia Bertozzi-Villa, Penelope Hancock, Punam Amratia, Katherine E. Battle, Ewan Cameron, Peter W. Gething, Daniel J. Weiss

## Abstract

As malaria incidence decreases and more countries move towards elimination, maps of malaria risk in low prevalence areas are increasingly needed. For low burden areas, disaggregation regression models have been developed to estimate risk at high spatial resolution from routine surveillance reports aggregated by administrative unit polygons. However, in areas with both routine surveillance data and prevalence surveys, models that make use of the spatial information from prevalence point-surveys have great potential. Using case studies in Indonesia, Senegal and Madagascar, we compare two methods for incorporating point-level, spatial information into disaggregation regression models. The first simply fits a Gaussian random field to prevalence point-surveys to create a new covariate. The second is a multi-likelihood model that is fitted jointly to prevalence point-surveys and polygon incidence data. We find that the simple model generally performs better than a baseline disaggregation model while the joint model performance was mixed. More generally, our results demonstrate that combining these types of data improves estimates of malaria incidence.

## Introduction

Global malaria incidence has decreased dramatically over the last 20 years (Bhatt et al., 2015; Weiss et al., 2019; Battle et al., 2019). This decrease has been accompanied by a strategic shift aiming for elimination in low incidence countries (World Health Organization, 2016; Newby et al., 2016). Accurate, high-resolution maps of malaria risk are vital in countries in the elimination and pre-elimination phases as they highlight the areas with ongoing *Plasmodium* transmission most in need of interventions (Sturrock et al., 2016; Cohen et al., 2017). Mapping malaria in low burden countries presents new challenges as traditional mapping of prevalence (Gething et al., 2011; Bhatt et al., 2017; Gething et al., 2012; Bhatt et al., 2015) using cluster-level surveys and model-based geostatistics are not necessarily effective in these areas (Sturrock et al., 2016, 2014). In low burden areas, very large sample sizes are needed before a prevalence survey is informative because so few individuals have detectable parasitaemia that most sample points will have no cases. In most cases, these large sample sizes are neither logistically nor financially feasible. However, the availability and quality of routine surveillance data of malaria case counts, typically aggregated by administrative unit polygons, is improving, thus providing an alternative data source for mapping malaria burden (Sturrock et al., 2016; Ohrt et al., 2015; Cibulskis et al., 2011). Advantageously, the routine surveillance data can be more sensitive than prevalence point-surveys in low transmission areas because the entire public health system is being used to passively monitor disease occurrence continually over a period of time (Cibulskis et al., 2011).

Disaggregation regression methods have been proposed as a way to model malaria burden using polygon-level, routine surveillance records of incidence (Sturrock et al., 2014; Wilson and Wakefield, 2018; Law et al., 2018; Taylor et al., 2017; Li et al., 2012; Johnson et al., 2019). Disaggregation regression requires an aggregation step in which the high-resolution estimates of disease incidence are summed to match the level of the administitive unit at which the incidence data are observed. An important consideration is whether the aggregation step occurs in link function space or in the response space. In the case of the identity link function, the two cases are the same (Moraga et al., 2017; Roksvåg et al., 2019; Wilson and Wakefield, 2018). However, when using a non-linear link function, the two cases imply very different models. In the case of the Normal– Poisson pairing with a log-link function, performing the aggregation step in the link space before transformation back to the response space produces a ‘geometric sum’ operation. This formulation has been used for computational convenience a number of times in the literature (Wang et al., 2018; Liu et al., 2011) but lacks the natural epidemiological interpretation provided by arithmetic summation in the response space.

The intent of this research is to assess the utility of extending disaggregation regression modelling approaches to map malaria incidence using both point- and polygon-level response data. In particular it focuses on two aspects where improvements in the disaggregation regression may be possible: (1) for modelling low-burden areas which have better coverage of prevalence surveys than polygon incidence data; and (2) for building the statistical relationships between the response (i.e. malaria incidence) and geospatial predictor data in heterogeneous landscapes. An ancillary benefit of a hybridized approach is that it will simultaneously produce estimates of both prevalence and incidence metrics, which may both be useful for policy makers (Cohen et al., 2017). Models are typically fitted to observations of one metric and then a secondary model is used to convert between prevalence and incidence *post hoc* (Battle et al., 2019; Bhatt et al., 2015), thus missing an opportunity to learn the relationship between prevalence and incidence at the same time as fitting the geographic model.

There are two broad ways that spatial information from prevalence surveys could be included in a dissaggregation regression model of incidence. Firstly, the information from prevalence surveys could be summarised using a separate model and then included as a covariate in the disaggregation model. If the model used to summarise the prevalence surveys was explicitly spatial, this approach would make the spatial information in the prevalence data available to the disaggregation model, thereby enhancing the ability to spatially disaggregate polygon-level cases within administrative units. However, this approach does not provide additional degrees of freedom in order to more accurately learn relationships between malaria risk and the environment, nor does it allow joint predictions of incidence and prevalence from a single model. This broad approach of summarising the information in a different data set using a separate model has previously been used in a number of contexts, including information on animal hosts (Shearer et al., 2016) or summarising temperature suitability for malaria parasites (Weiss et al., 2014b), which were subsequently used as inputs for modeling malaria prevalence (Bhatt et al., 2015; Weiss et al., 2019).

Fully combining observations of incidence and prevalence in a joint model, with multiple likelihoods, addresses the limitations of a simple model using a prevelance map as a covariate. Advantageously, as the additional malariometric data are being used as response data, they provide more degrees of freedom with which to learn relationships between malaria risk and the environment. Such a model can also learn the relationship between different types of malaria response metrics at the same time as making spatial estimates, thereby producing statistically and epidemiologically consistent outputs for both incidence and prevalence. While a joint model provides the opportunity to learn the relationship between prevalence and incidence, this is technically challenging as these two data types measure disease intensity on different scales. Point-surveys are a measurement of prevalence in the range [0, 1] that quantify parasite rate at a specific point in time. In contrast, routine surveillance measures incidence in the range [0, ∞] over a longer period of time (e.g., a year) during which individuals can have multiple malaria infections. The case of using areal and point data together with different likelihoods and different link functions has been examined previously (Wang et al., 2018) but has required that the aggregation step be performed in the link function space. Disaggregation regression models in which the aggregation step is performed in the natural response space have been examined (Wilson and Wakefield, 2018; Taylor et al., 2017), but without combining point data with areal data or using dual likelihoods for multi-metric data.

Here we compare two methods for using spatial information from prevalence surveys to inform a disaggregation model fitted to polygon incidence data of *Plasmodium falciparum* malaria. The first, simpler, model summarises the spatial information in the prevalence point-surveys by fitting a spatial Gaussian process model to the surveys. Predictions from this model are then used as a covariate in the disaggregation model. Secondly, we formulate a joint model that combines polygon incidence data and prevalence point-surveys using separate likelihoods for both data types. We relate the differing malariometric measures by using a previously estimated relationship within the model (Cameron et al., 2015) which is then adjusted as part of the model fitting process. Unlike previous studies, this model combines areal and point level data, with different likelihoods, without performing the aggregation step in the link function space. We then compare results from the two models with those made using a polygon-only, disaggregation model similar to previous models (Sturrock et al., 2014; Wilson and Wakefield, 2018). All models are fitted to data from Indonesia, Senegal and Madagascar to provide a set of case studies from disparate geographic settings and with differing levels of malaria endemicity.

## Materials and methods

### Malaria data

We used two data sources that quantify malaria burden: prevalence point-surveys and polygon incidence data. Prevalence point-surveys consist of geo-located survey clusters wherein all sampled individuals are tested for malaria and the positive cases as well as the total number of children tested is recorded. Polygon incidence data is aggregated to administrative units (e.g. districts or provinces) summarizing data reported from hospitals and health facilities. Unlike the point data, polygon-level reports only include numbers of cases and not the numbers of individuals in each administrative unit. As such, to determine an incidence rate we rely on gridded population surfaces, summarised to administrative unit boundaries, to provide the denominator. The prevalence point-survey data were extracted from the Malaria Atlas Project database (Bhatt et al., 2015; Guerra et al., 2007; Pfeffer et al., 2018). As the prevalence point-surveys cover different age ranges they were standardised to the 2–10 year-range using a previously published model (Smith et al., 2007). As described, the age standardisation model gives the surveys with zero positive cases a small positive prevalence. The polygon incidence data were collated from various government reports and adjusted for incompleteness using methods defined by Cibulskis and colleagues (Cibulskis et al., 2011; Weiss et al., 2019). These adjustments account for underreporting of clinical cases due to lack of treatment seeking, missing case reports (from a health facility that reported for 11 months in a year for example), and cases that sought medical attention outside the public health systems (Battle et al., 2016). Where species specific reports were given, these were used, and in reports that did not distinguish between species of *Plasmodium* the national estimate of the ratio between *P. falciparum* and *Plasmodium vivax* cases was used to estimate numbers of *P. falciparum* cases specifically. These adjustments were uniform across each country. The polygon incidence data can be seen in Panel A of Figures 1–3.

**Fig. 1.**
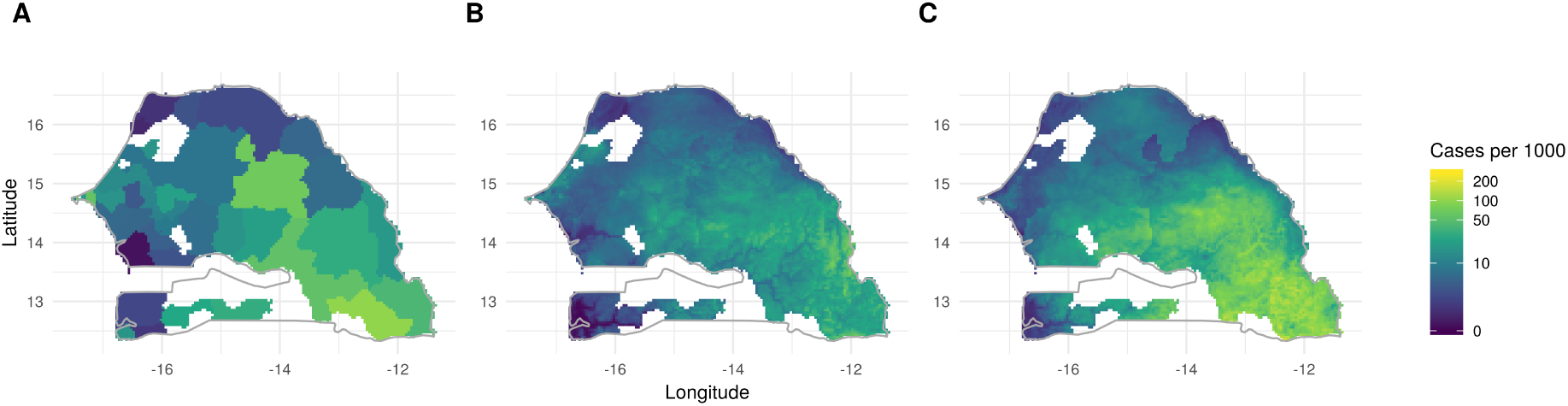
Reported incidence data and modelled incidence maps for Senegal. The national boundary of Senegal is shown in grey and missing data is left white. The adjusted input aggregated data is plotted in Panel A, while Panel B maps the predictions of the prevalence Gaussian Process model for for spatially cross-validated out-of-sample polygons and Panel C maps the predicted incidence from the joint model.

**Fig. 2.**
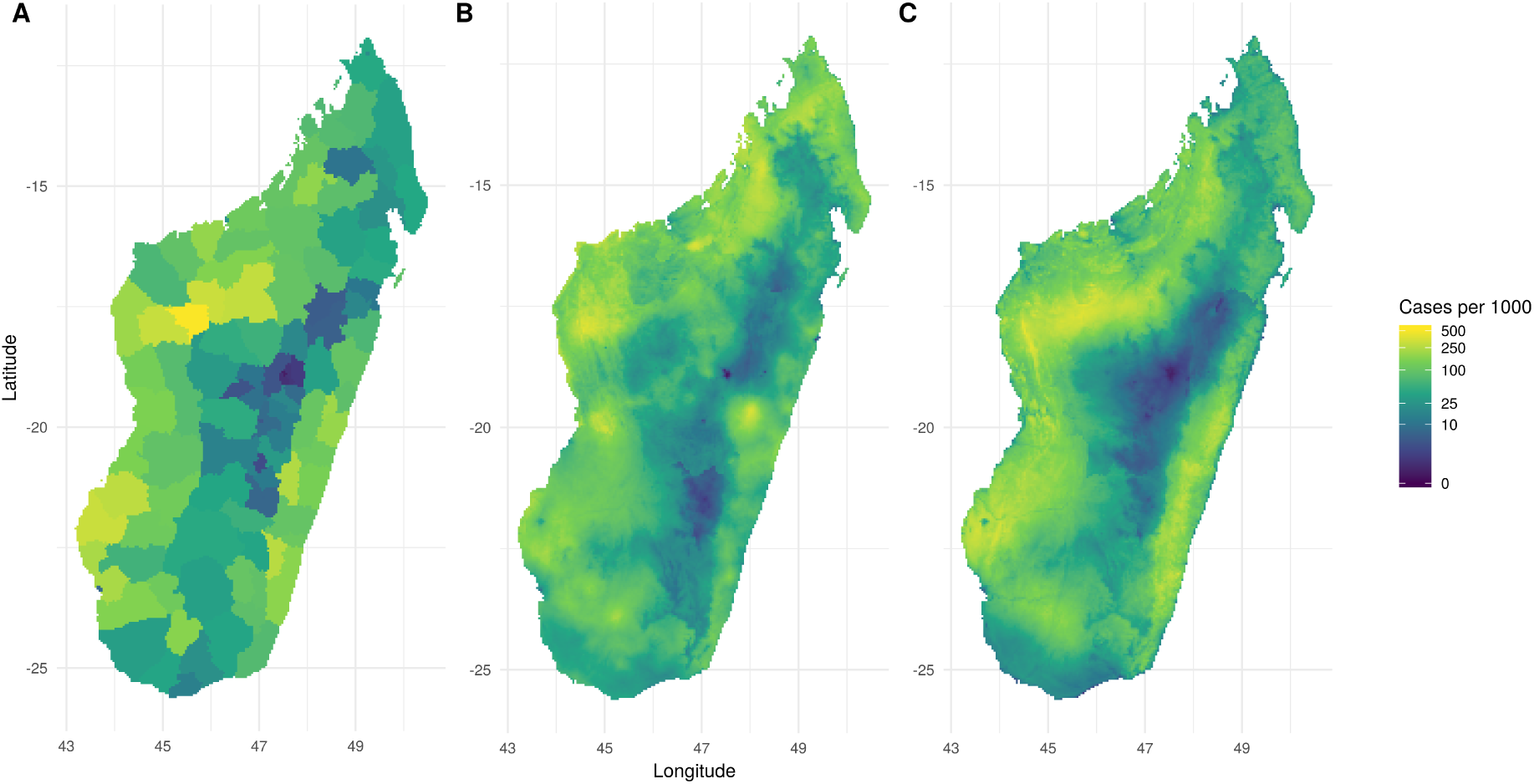
Reported incidence data and modelled incidence maps for Madagascar. The adjusted input aggregated data is plotted in Panel A, while Panel B maps the predictions of the prevalence Gaussian Process model for for spatially cross-validated out-of-sample polygons and Panel C maps the predicted incidence from the joint model.

**Fig. 3.**
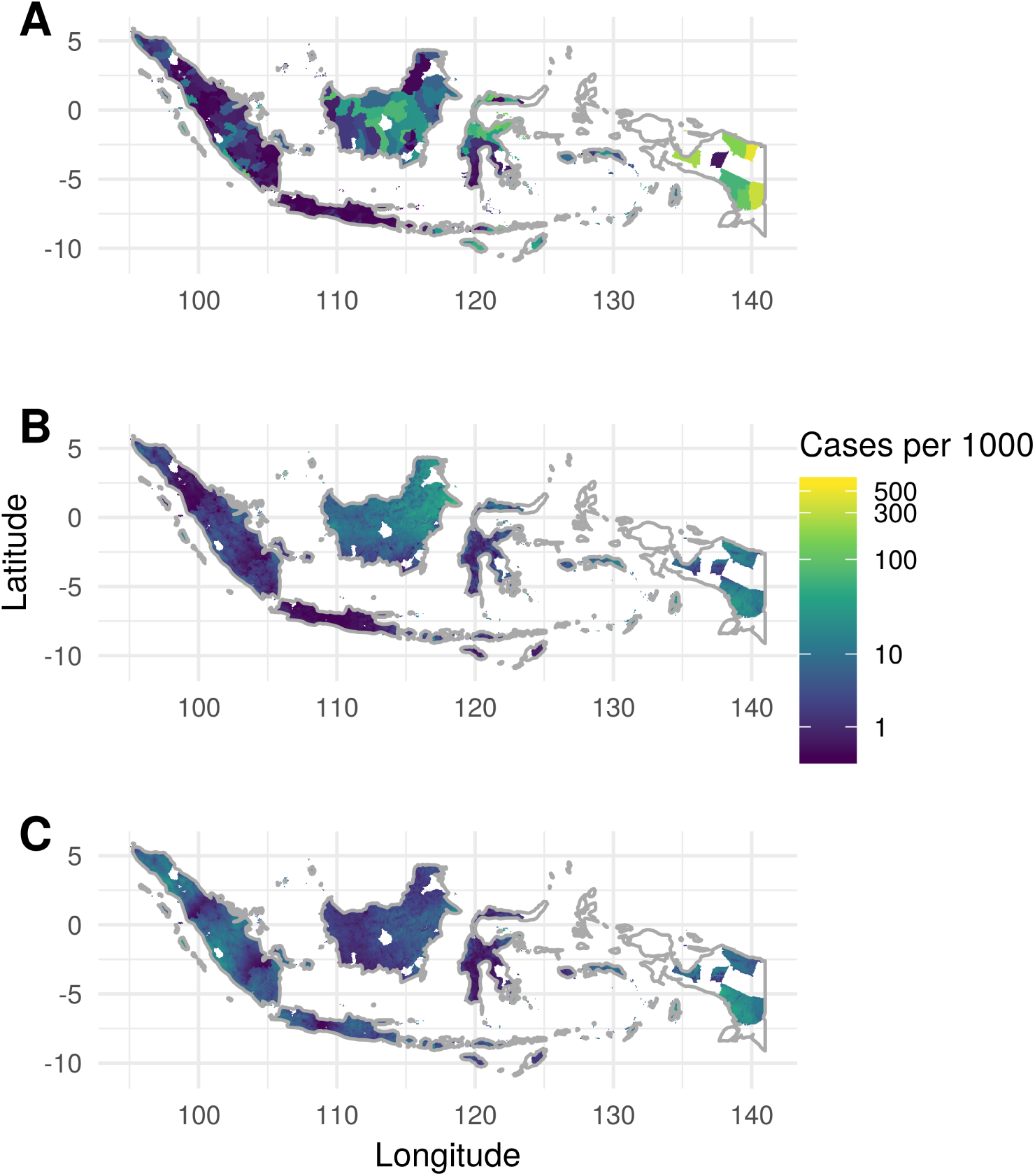
Reported incidence data and modelled incidence maps for Indonesia. The national boundary of Indonesia is shown in grey and missing data is left white. The adjusted input aggregated data is plotted in Panel A, while Panel B maps the predictions of the prevalence Gaussian Process model for for spatially cross-validated out-of-sample polygons and Panel C maps the predicted incidence from the joint model.

We selected Indonesia, Senegal and Madagascar as case examples as they all have abundant subnational surveillance data and country-wide surveys from approximately the same periods. To minimise temporal effects we selected one year of polygon incidence data and the surrounding five years of prevalence point-survey data for each country. Within this five year period, we considered malaria unchanging and did not model time explicitely. For Indonesia we selected polygon incidence data from 2012 that covers 379 administrative units, and prevalence data from 2010 to 2014 that consists of 1,233 survey clusters (i.e. unique locations), representing 230,747 individuals. For Senegal we selected 2015 for polygon incidence data (41 administrative units) and 2013 to 2017 for prevalence data (804 clusters, 17,037 individuals). Finally, for Madagascar we selected 2013 for polygon incidence (110 administrative units) and 2011 to 2015 for prevalence data (1,049 clusters, 36,411 individuals).

### Population data

Raster surfaces of population for the years 2005, 2010 and 2015, were created using a hybrid mosaic of data from the Gridded Population of the World v4 (NASA, 2018) and WorldPop (Tatem, 2017), with the latter taking priority for those pixels where both sources had population data. For each year, the interpolated population surfaces were adjusted to match national population estimates from the UN. Finally, the population surfaces were masked by environmental suitability so that only populations at risk were included (Weiss et al., 2019).

### Covariate data

We considered a suite of environmental and anthropological covariates, at a resolution of approximately 5 *×* 5 kilometres at the equator that included land surface temperature annual mean and standard deviation, enhanced vegetation index (EVI), *P. falciparum* temperature suitability index (Weiss et al., 2014b), elevation (NASA LP DAAC, 2013), tassel cap brightness, tassel cap wetness, accessibility to cities (Weiss et al., 2018), night lights (Elvidge et al., 2017) and proportion of urban land cover (Esch et al., 2018). The land surface temperature, EVI, and tasseled cap indices were derived from satellite imagery and gap-filled to remove missing data caused by factors like cloud-cover (Weiss et al., 2014a) and rescaled to a spatial resolution of approximately 5 *×* 5km (Weiss et al., 2015) that defined the output of the final prevalence and incidence maps. Some covariates were log-transformed to remove skewness or removed due to multicollinearity with other predictor variables using the threshold of 0.8. The covariates were standardised to have a mean of zero and a standard deviation of one.

### Baseline Disaggregation Model

Values at the aggregated, polygon level are given the subscript *a* while pixel or point level variables are indexed with *b*. The polygon incidence case count data, *y*_*a*_ is given a Poisson likelihood

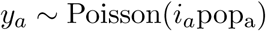

where *i*_*a*_ is the estimated polygon incidence rate and pop_a_ is the population at risk within that admin unit polygon (as apposed to the true health centre catchment area).

Incidence rate is linked to latent pixel-level incidence (*i*_*b*_), prevalence (*p*_*b*_) and predictor variables by the following system of equations.

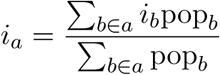

Here, *b* ∈*a* denotes that the summation is over the pixels in polygon *a*. Incidence is related to prevalence by

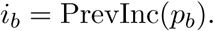

Here PrevInc is a function from a previously fitted model (Cameron et al., 2015)

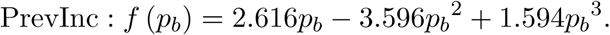

The linear predictor of the model, *η*_*b*_, is related to the latent prevalence scale by a typical logit link function.

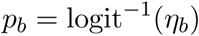

The form of this set of link functions means we calculated predictions of prevalence and incidence simultaneously whether both data types or just one were used.

The linear predictor is composed of an intercept, *b*_0_, covariates, *X*, and a vector of regression coefficients ***β***. We also include a spatial, Gaussian random field, *u*_*s*_(*ρ, σ*_*u*_) and a polygon-level iid random effect, *v*_*a*_(*σ*_*v*_).

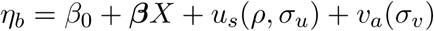

The Gaussian spatial effect *u*(*s, ρ, σ*_*u*_) has a Matérn covariance function and two hyper parameters: *ρ*, the nominal range on the longitude-latitude scale (beyond which correlation is *<* 0.1) and *σ*_*u*_, the marginal standard deviation. The iid random effect, *v*_*a*_ ∼Normal(0, *σ*_*v*_), was grouped by polygon, with all pixels within polygon *j* being grouped together. Internally, this effect is parameterised as the log of the precision, 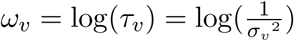 to improve numeric stability. This random effect modelled both missing covariates and extra-Poisson sampling error.

Finally, we complete the model by setting priors on the parameters *β*_0_, ***β***, *ρ, σ*_*u*_ and *σ*_*v*_. The intercept was given a wide prior, *b*_0_ ∼Normal(−2, 4), with a mean relating to a prevalence of 0.12 as we know *a priori* that these countries have low or medium levels of malaria transmission. We set independent, regularising priors on the regression coefficients *β*_*i*_ ∼Normal(0, 0.04). Given the standardised covariates, an intercept of −3 and a regression coefficient from the 95% interquartile range of this distribution, each covariate would be able to predict prevalences between 0.004 and 0.27. This prior encodes our belief that the full range of malaria transmission can not be explained by a single covariate and our desire to regularise the model. This regularisation is particularly important given the small number of administrative units in Senegal (n = 46) and Madagascar (n = 110).

We assigned *ρ* and *σ*_*u*_ a joint penalised complexity prior (Fuglstad et al., 2018) such that *P* (*ρ < ζ*) = 0.00001 and *P* (*σ*_*u*_ *> ξ*) = 0.00001. We used different *ζ* and *ξ* values for each country: Indonesia *ζ* = 3, *ξ* = 1, Senegal *ζ* = 1, *ξ* = 0.5 and Madagascar *ζ* = 1, *ξ* = 1. We believe that a large proportion of the variance of malaria prevalence and incidence cannot be explained by a linear combination of the covariates selected at the scale of individual countries (Bhatt et al., 2017), so we set this prior such that the random field could explain most of the range of the data. As Senegal has a lower range of incidences in the data we set *ξ* to a smaller value for this country.

We assigned *σ*_*v*_ a penalised complexity prior (Simpson et al., 2017) such that *P* (*σ*_*v*_ *>* 0.05) = 0.0000001. This was based on a comparison of the variance of Poisson random variables, with rates given by the number of cases observed, and an separately derived upper and lower bound for the case counts using the approach defined by Cibulskis and colleagues (Cibulskis et al., 2011). We found that an iid effect with a standard deviation of 0.05 was able to account for the discrepancy between the assumed Poisson error and the separately derived measurement error.

The models were implemented and fitted in R (R Core Team, 2018) using Template Model Builder (Kristensen et al., 2016) which allows a Laplace approximation of the posterior to be calculated. We note that R-INLA (Lindgren and Rue, 2015) can be used to fit disaggregation models but only when a linear link function is being used (Wilson and Wakefield, 2018). The hyperparameters are fitted using empirical Bayes whereby the hyperparameters are learned from the data but are treated as point estimates rather than using the full posterior of the hyperparameters.

### Prevalence Gaussian process covariate model

The prevalence Gaussian Process model (henceforth the prevalence GP model) is the same as the baseline disaggregation model except that it has one extra covariate. This covariate is created by fitting a Gaussian random field to the prevalence survey data. For each country we fitted a binomial likelihood, hierarchical Gaussian random field with the same hyperpriors for *ρ* and *σ*_*u*_ as above. These models were fitted using R-INLA (Lindgren and Rue, 2015). To be in the correct scale for the dissagregation model, the inverse logit of the predicted Gaussian field (i.e. the linear predictor of the model) was used as the additional covariate.

### Full joint model

The final model is a joint-likelihood model with separate likelihoods for prevalence point-surveys and polygon incidence data. The polygon data are assigned a Poisson likelihood as before. Additionally, the point-survey data, with positive cases *z*_*b*_, are given a binomial likelihood

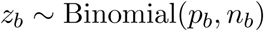

where *p*_*b*_ is the estimated prevalence and *n*_*b*_ is the observed survey sample size. As this model has both prevalence and incidence data we add a parameter *α* that modifies the relationship between the two.

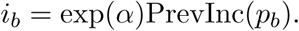

The only further additions to the baseline model are in the linear predictor which becomes

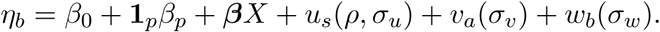

As well as the global intercept, *β*_0_, this model has a prevalence survey specific intercept *β*_*p*_ where the indicator function, **1**_*p*_ denotes that this term is zero except when a prevalence point-survey is being considered. The iid random effect, *v*_*a*_ ∼Norm(0, *σ*_*v*_), was again grouped by polygon, with all pixels and point-surveys within polygon *a* being in the same group as polygon *a*. The second iid random effect, *w*_*b*_ ∼ Normal(0, *σ*_*w*_), was applied to each point-survey. To improve numeric stability this effect is also parameterised internally as the log of the precision, 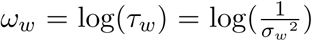. This effect modelled extra-binomial sampling noise. As such, this random effect is not included in the predicted uncertainty in the incidence or prevalence layers.

We assigned *σ*_*w*_ a penalised complexity prior such that *P* (*σ*_*w*_ *> ϕ*) = 0.0000001. This was chosen by finding the maximum difference in prevalence between point-surveys (with a sample size greater than 500 individuals) within the same raster pixel. The differences between points within the same pixel can only be accounted for by the binomial error and this iid effect. Given that the error on a prevalence estimate with sample size greater than 500 is quite small, the iid effect needs to be able to explain this difference. In Senegal and Madagascar this value was relatively small so we set *ϕ* = 0.05. In Indonesia however, there was a high density of prevalence surveys and heterogeneity in estimated prevalence within single pixels. Therefore we set *ϕ* = 0.3.

Given that the PrevInc relationship is fitted to the best available data, we have fairly strong *a priori* confidence in it. Therefore, our prior belief is that exp(*α*) is close to one (i.e. the relationship remains unchanged) and therefore that *α* is close to zero. We set our prior as *α* ∼ Normal(0, 0.001).

### Experiments

To compare the three models we used two cross-validation schemes. In the first (random), the incidence data was split into ten cross-validation folds while all the prevalence data was used in each case (Figure S1. In the second validation scheme, the incidence data was split into spatial cross-validation folds, using k means clustering on polygon centroids, while again all prevalence points were used in all folds (Figure S2). The number of folds was seven for Indonesia, five for Senegal and three for Madagascar due to their differing sizes and epidemiological settings. This scheme tests specifically whether the joint model can improve predictions by increasing geographic data coverage.

We considered the ability of the model to predict polygon incidence to be our main objective and our performance metric for this was mean absolute error (MAE). As the models were fitted on data on different scales we found that observations and predictions were sometimes correlated but shifted from the one-one line (i.e. were biased) and therefore correlation metrics were misleading. To assess how well the models were calibrated we considered coverage of the 80% predictive credible intervals on the hold-out data.

## Results

Under the random cross-validation scheme, the prevalence GP model performed best in Senegal and Madagascar while the joint model performed best in Indonesia (Table 1). The differences were relatively small in all three countries. This lack of strong differences is highlighted by there being no clear differences in scatter plots of observed and predicted data across the three methods (Figure 4).

**Table 1.**
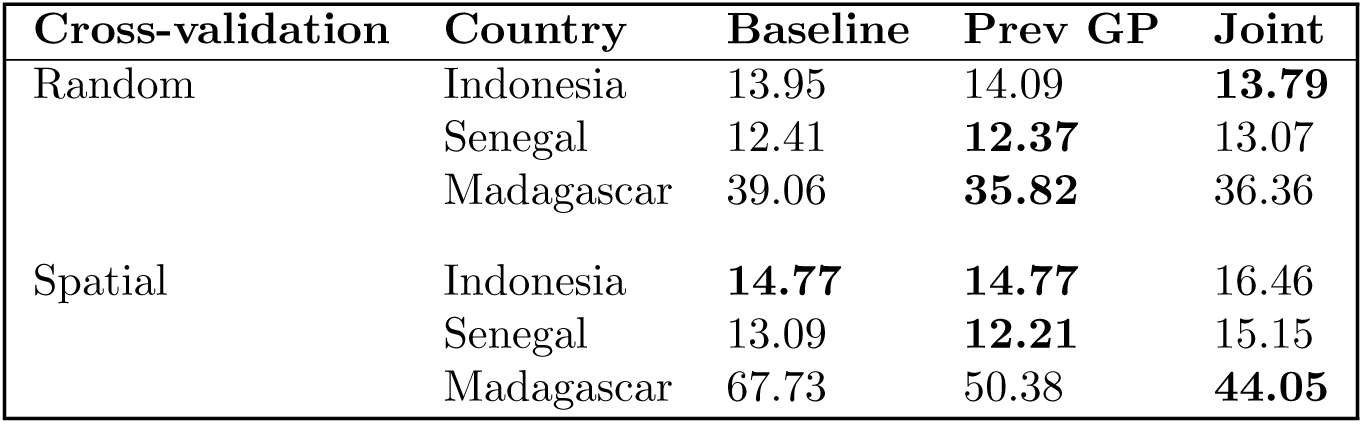
Summary of out-of-sample accuracy for all cross-validation experiments. Mean absolute error of predicted incidence rate against out-of-sample observed data for three countries.

| Cross-validation | Country | Baseline | Prev GP | Joint |
| --- | --- | --- | --- | --- |
| Random | Indonesia | 13.95 | 14.09 | **13.79** |
|  | Senegal | 12.41 | **12.37** | 13.07 |
|  | Madagascar | 39.06 | **35.82** | 36.36 |
| Spatial | Indonesia | **14.77** | **14.77** | 16.46 |
|  | Senegal | 13.09 | **12.21** | 15.15 |
|  | Madagascar | 67.73 | 50.38 | **44.05** |

**Fig. 4.**
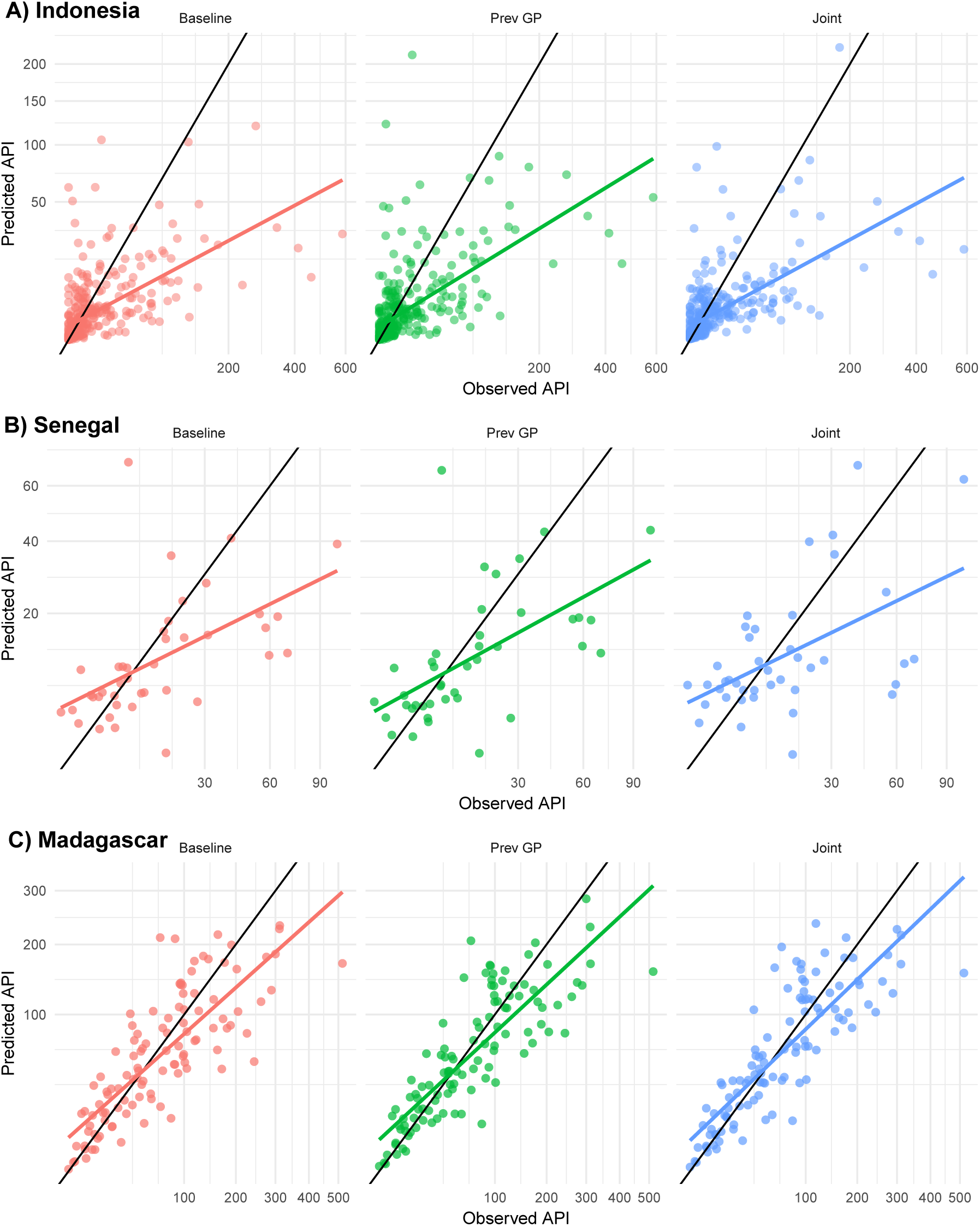
Observed-predicted plots (square root scale) of modelled annual malaria incidence (cases per 1000) by country from the random cross-validation experiments for Indonesia (Panel A), Senegal (Panel B) and Madagascar (Panel C). Results from the baseline disaggregation model are shown in red, the prevalence GP model is shown in green while the joint model is shown in blue. The one-one line is shown with a black line and a simple linear regression through the points is shown by a coloured line.

Under the spatial cross-validation scheme, the baseline model and prevalence GP models performed best in Indonesia, the prevalence GP performed best in Senegal while the joint model performed best in Madagascar (Table 1). In contrast to the random cross-validation results, the differences between models was quite strong. Furthermore, notable differences can be seen in the scatter plots of observed and predicted values (Figure 5). In Indonesia it can be seen that the joint model is more strongly biased at low incidence values with many data points being overpredicted. However, the joint model clearly performs better in Madagascar with the polygon-only model unable to predict high incidence observations accurately. Out-of-sample predictions, under spatial cross validation, from the prevalence GP model and full joint model can be seen in Figures 1 – 3.

**Fig. 5.**
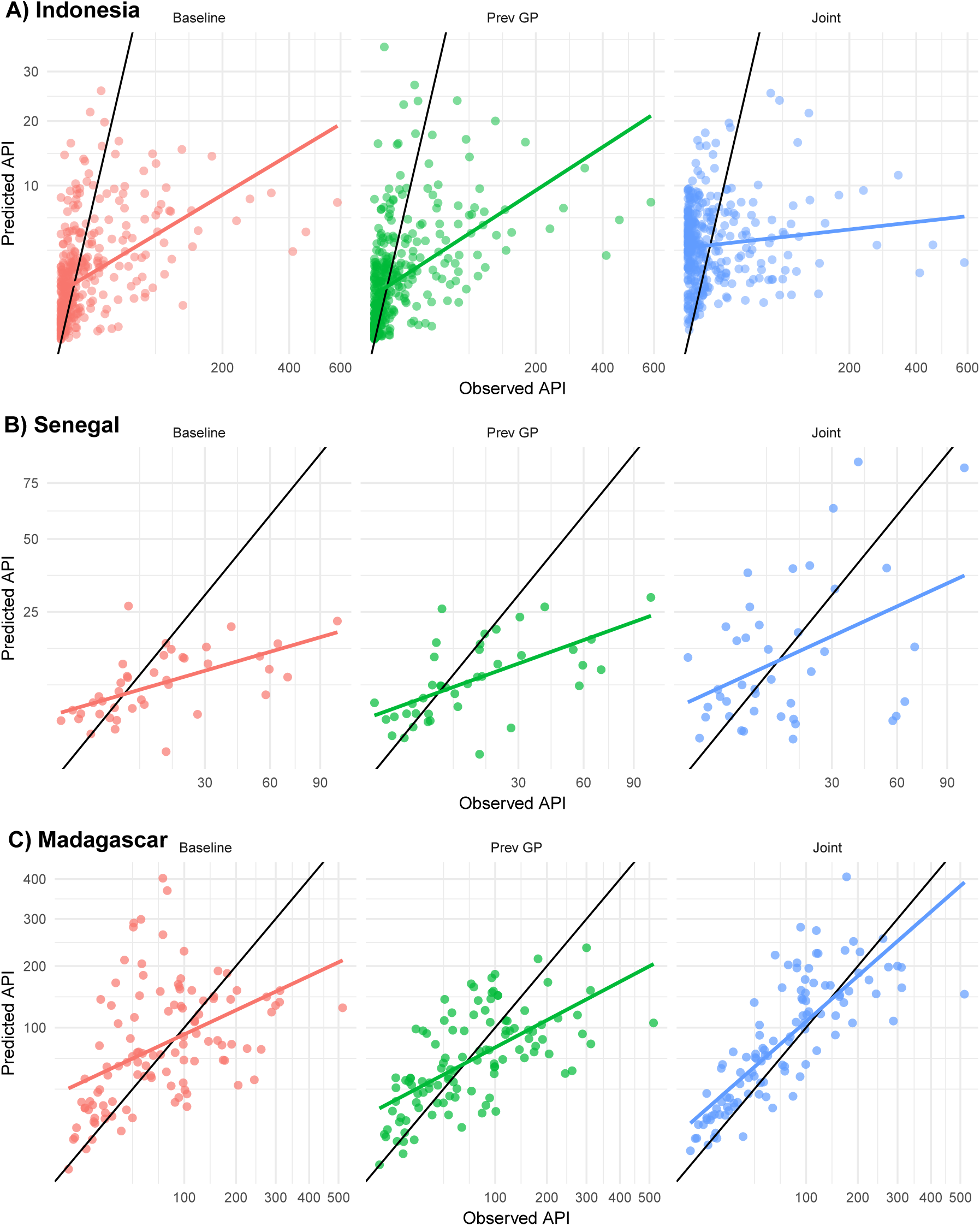
Observed-predicted plots (square root scale) of modelled annual malaria incidence (cases per 1000) by country from the spatial cross-validation experiments for Indonesia (Panel A), Senegal (Panel B) and Madagascar (Panel C). Results from the baseline disaggregation model are shown in red, the prevalence GP model is shown in green while the joint model is shown in blue. The one-one line is shown with a black line and a simple linear regression through the points is shown by a coloured line.

All models seem to be fairly well calibrated (Table 2). The proportion of out-of-sample incidence datapoints being within their 80% credible intervals ranged between 0.51 and 0.88. However, in most cases coverage was between 0.7 and 0.8 implying that the models were a little overconfident in their predictions. There was no clear difference in calibration between the different models.

**Table 2.** Summary of coverage of 80% credible intervals. The proportion of held out data points that fall within their 80% credible intervals. Cases where this is below 0.7 are highlighted in bold.

| Cross-validation | Country | Baseline | Prev GP | Joint |
| --- | --- | --- | --- | --- |
| Random | Indonesia | 0.73 | 0.72 | 0.72 |
|  | Senegal | 0.76 | 0.78 | 0.80 |
|  | Madagascar | 0.78 | 0.79 | 0.78 |
| Spatial | Indonesia | 0.71 | 0.72 | **0.51** |
|  | Senegal | 0.78 | 0.88 | 0.71 |
|  | Madagascar | **0.67** | 0.70 | 0.72 |

We can further investigate why the models performed as they did by examining the parameters estimated in the models fitted to all data (Tables S1–S3). Firstly we can compare the regression parameter for the prevalence GP covariate in the three countries noting that in Indonesia the prevalence GP model performed worse than baseline under random cross-validation and had equal performance to baseline under spatial cross-validation. We see that the regression parameter for this covariate was small in Indonesia (mean = 0.06, sd = 0.12) but relatively large and positive in both Senegal (mean = 0.30, sd = 0.17) and Madagascar (mean = 0.36, sd = 0.07).

In Madagascar the joint model performed the best in the spatial cross-validation scheme and better than the baseline in the random cross-validation scheme. Comparing the estimated parameters of the joint model between Madagascar and the other two countries therefore is useful. The prevalence intercept, *β*_*p*_, is large in the Senegal fit (mean = 1.36, sd = 0.13) but small in Indonesia (mean = 0.03, sd = 0.20) and Madagascar (mean = 0.07, sd = 0.10). This implies there is a strong discrepency (given the prevalence to incidence model) between the prevalence and incidence data in Senegal. Furthermore, the standard deviation of the prevalence point iid effect, *w*_*b*_(*σ*_*w*_), is much larger in Indonesia (mean *ω*_*w*_ = −2.6, sd *ω*_*w*_ = 0.09 which corresponds to a mean of *σ*_*w*_ of 13.46) than in Senegal (mean *ω*_*w*_ = −1.03, sd *ω*_*w*_ = 0.13 which corresponds to a mean of *σ*_*w*_ of 2.80) or Madagascar (mean *ω*_*w*_ = −0.77, sd *ω*_*w*_ = 0.11 which corresponds to a mean of *σ*_*w*_ of 2.16). This implies there is a lot of noise in the prevalence data in Indonesia.

We set a strong prior on *α* being close to one, encoding our belief that the incidence prevalence relationship should be close to the previously fitted model. The estimated value for *α* in all three countries is very close to one (Tables S1–S3). While this might be driven by the prior, we can conclude that there is no strong evidence from the data that this relationship should be scaled differently by country.

Overall, inclusion of the spatial information from prevalence surveys yielded predictions that were as good or better than the baseline model in all six experiments (three countries and two cross-validation schemes). The prevalence GP model was as good or better than baseline in five out of six experiments. In contrast, the joint model was only better than baseline in three out of six experiments.

## Discussion

We have compared the predictive performance of three models: a baseline polygon-only model; a disaggregation model with spatial information from prevalence surveys included as an additional covariate from a separate Gaussian process (GP) model; and a model that jointly learns from polygon incidence data and prevalence point-surveys. Overall the prevalence GP model appeared to perform best. While the joint model sometimes performed best it also performed worse than baseline in half of the experiments. Therefore, fitting a spatial Gaussian process to prevalence points and including these predictions seems to be a more reliable way of using spatial information from prevalence points. However, given that this comparison was conducted on datasets from only three countries, it is challenging to draw firm conclusions.

A full joint model using both prevalence surveys and incidence data gains a large number of additional degrees of freedom compared to the baseline or prevalence GP models. Therefore, it is worth considering why the performance of this model was generally less good than the simpler prevalence GP model that did not benefit from the additional degrees of freedom. One potential reason is that the malariometric data are on different scales. Here we have used a previously fitted model (Cameron et al., 2015) to inform the joint model. However, this model was calibrated using relatively few matched prevalence and incidence surveys as few of these have been conducted and published. Although we added the parameter *α* that scales this relationship, it is a very simple scaling. Furthermore the true relationship between prevalence and incidence is likely to vary spatially as aspects such as immunity, seasonality, and population age-structure are not constant (Cameron et al., 2015; Battle et al., 2015; Reiner et al., 2015). In using a joint model we are accepting these limitations in the hope that the benefits of including additional data outweigh the costs of using mismatched data.

Future models could potentially be improved by using a more flexible approach for addressing the shortcomings of the prevalence-incidence relationship (Cameron et al., 2015) being used in this context. This could be by estimating the parameters of the polynomial jointly with the rest of the model. Informative priors based on the original model could be used to regularise this joint fit both to prevent improbable inferences but also because if the relationship were too flexible, the information from the prevalence data might not contribute to informing the regression parameters and spatial random field. This is particularly true for model forms such as a spline or a Gaussian process on the relationship between prevalence and incidence. For the model to handle noisy or biased prevalence point-surveys, the modeller can control the iid random effect on the point-surveys, *w*_*b*_ and the prevalence intercept *β*_*p*_. Here we have tried to maximise the influence of the prevalence data by setting the prior based on the belief that the random effect should only explain extra-binomial variation that is impossible to derive from the covariates (e.g. based on the differences in prevalence surveys within the same pixel). Weakening this prior will allow the iid effect to explain more of the prevalence point-survey variation which both reduces the potential statistical power gained by adding the point-surveys but also reduces the effects of biased or noisy estimates.

In this research we have used only linear covariates but previous work has demonstrated that simple linear combinations of environmental covariates cannot fully explain malaria risk (Bhatt et al., 2017). A number of methods could be used to include non-linear effects of covariates and interactions into the model. Firstly, machine learning models could be fitted to the prevalence data and then predictions from these models could be used as covariates in the full model (Bhatt et al., 2017). This approach is feasible but would not allow any information from the polygons to inform non-linear relationships. Directly modelling non-linear effects in the full model could be achieved by including simple non-linear functions such as splines (Sissoko et al., 2017; Sewe et al., 2017; Hundessa et al., 2018), though the increased model complexity would require more data than was used in Senegal and Madagascar in this study. Finally, Gaussian process regression, with smoothly varying effects in environmental and geographic space could be used (Law et al., 2018). Unfortunately, each of these options is computationally expensive without variational Bayes or other approximations (Law et al., 2018; Ton et al., 2018), which can be difficult to derive. Additionally these models require a large volume of response data and careful regularisation for good predictive performance.

We used three case studies, limited by the number of countries with good aggregated incidence data as well as good prevalence survey data. Given the small number of case studies it is hard to determine when these methods are likely to be most effective. However, the greatest benefits here were seen in Madagascar, a country with more intermediate transmission intensities. In the future, two groups of countries might particularly benefit from the methods presented here. Firstly countries who have had large prevalence surveys in the past and whose reporting systems are improving, such as Ethiopia, might benefit from these methods. Secondly, countries that have lots of prevalence surveys and are adjacent to countries with good reporting systems, (e.g. Papua New Guinea and neighbouring Indonesia), might also benefit from models that share information between countries.

## Conclusion

Overall, we have shown that including spatial information from prevalence surveys generally improves the predictive performance of disaggregation regression of aggregated incidence data. However, we found that the more complex joint model was unreliable in its predictive performance. In contrast, summarising the spatial information from the prevalence surveys by fitting a spatial Gaussian process model and using predictions from this model as a new covariate nearly always improved predictive performance. As more countries produce reliable routine surveillance data, and as more countries reduce their malaria prevalences to the point where prevalence surveys are no longer sensitive, disaggregation regression will become more commonly used. Methods such as those presented here should be utilized and further refined to improve disaggregation regression results where and when the requisite data are available.

## Data Availability

The data is used under various data agreements so cannot be shared. Reproducible code with dummy data is available on request and will be hosted on GitHub soon.

## Supporting information

## Acknowledgments

The authors acknowledge the National Malaria Control Programme of Madagascar for sharing their routine case data for this analysis.

## Notes

### Competing Interest Statement

The authors have declared no competing interest.

### Funding Statement

Gates Foundation

